# Development of the Centralized Interactive Phenomics Resource (CIPHER) Standard for Electronic Health Data-Based Phenomics Knowledgebase

**DOI:** 10.1101/2022.09.12.22279792

**Authors:** Jacqueline Honerlaw, Yuk-Lam Ho, Francesca Fontin, Jeffrey Gosian, Monika Maripuri, Michael Murray, Rahul Sangar, Ashley Galloway, Andrew J. Zimolzak, Stacey B. Whitbourne, Juan P. Casas, Rachel B. Ramoni, David R. Gagnon, Tianxi Cai, Katherine P. Liao, J. Michael Gaziano, Sumitra Muralidhar, Kelly Cho

**Affiliations:** Massachusetts Veterans Epidemiology Research and Information Center (MAVERIC), VA Boston Healthcare System, Boston, MA, USA; Center for Innovations in Quality, Effectiveness and Safety, Michael E. DeBakey VA Medical Center, Houston, TX, USA; Department of Medicine, Baylor College of Medicine, Houston, TX, USA; Department of Medicine, Harvard Medical School, Boston, MA, USA; Department of Medicine, Division of Aging, Brigham and Women’s Hospital, Boston, MA, USA; Office of Research and Development, Veterans Health Administration, Washington, DC, USA; School of Public Health, Department of Biostatistics, Boston University, Boston, MA, USA; Department of Biostatistics, Harvard T. H. Chan School of Public Health, Boston, MA, USA; Department of Biomedical Informatics, Harvard Medical School, Boston, MA, USA; Medicine, Rheumatology, VA Boston Healthcare System, Boston, MA, USA; Division of Rheumatology, Inflammation, and Immunity, Brigham and Women’s Hospital, Boston, MA, USA; Department of Medicine & Biomedical Informatics, Harvard Medical School, Boston, MA, USA

## Abstract

The development of phenotypes using electronic health records is a resource intensive process. Therefore, the cataloging of phenotype algorithm metadata for reuse is critical to accelerate clinical research. The Department of Veterans Affairs Office of Research and Development has developed a phenomics knowledgebase library, CIPHER (Centralized Interactive Phenomics Research), which improves upon existing phenomics library models to help advance innovation in clinical research by using the CIPHER phenotype collection standard. The CIPHER standard was iteratively developed with phenomics experts and has been used to capture over 5,000 phenotypes. We describe the development of the CIPHER standard for phenotype metadata collection, its current application to the largest healthcare system in the United States, and the future expansion of the CIPHER knowledgebase as a public resource for phenotyping.

## INTRODUCTION

Electronic health records (EHR) are routinely leveraged to generate phenotypes for use in clinical research and healthcare operations. The Department of Veterans Affairs (VA) EHR system supports the largest healthcare network in the United States, consisting of over 1,290 facilities and 6 million Veterans receiving care annually, totaling 24 million users in the last 20 years.^1^ The VA EHR contains a wide breadth and depth of data from outpatient and inpatient settings, including unique content for the Veteran population such as service-related benefits and screening for military environmental exposures. The availability of over 20 years of structured and unstructured data has provided a valuable asset for VA research.^2^ Linkage with internal datasets (including patient registries and clinical trials) and external datasets such as Department of Defense, Centers for Medicare and Medicaid Services and the National Death Index supplement the VA EHR to provide a more complete picture of Veteran health. Knowledge extracted from such rich data sources can also greatly benefit EHR research and downstream clinical studies at large.

Expertise in both phenomics science and the intricacies of VA data are needed to develop EHR-based phenotypes. For example, multiple definitions of binary post-traumatic stress disorder (PTSD) status have been identified using the frequency, source (such as mental health provider) and location (inpatient or outpatient) of PTSD diagnosis codes. Harrington and colleagues built upon this work by developing a model to predict the probability of having PTSD, providing a flexible and adaptable definition for future applications.^3^ Understanding of patient encounters within the health system, patterns of code usage, expertise with curating and processing large datasets, and validation of algorithms are required to accurately define complex phenotypes such as PTSD. Given the time and resources required to develop phenotypes, the resulting algorithm metadata need to be captured and made available for future use and application, which is rarely done^4-6^. To meet these needs across the national healthcare system, VA has developed a phenomics knowledgebase library, Centralized Interactive Phenomics Resources (CIPHER), which enables reusability of EHR-based algorithms and increases efficiency and innovation in phenomics. (Figure 1)

**Figure 1:**
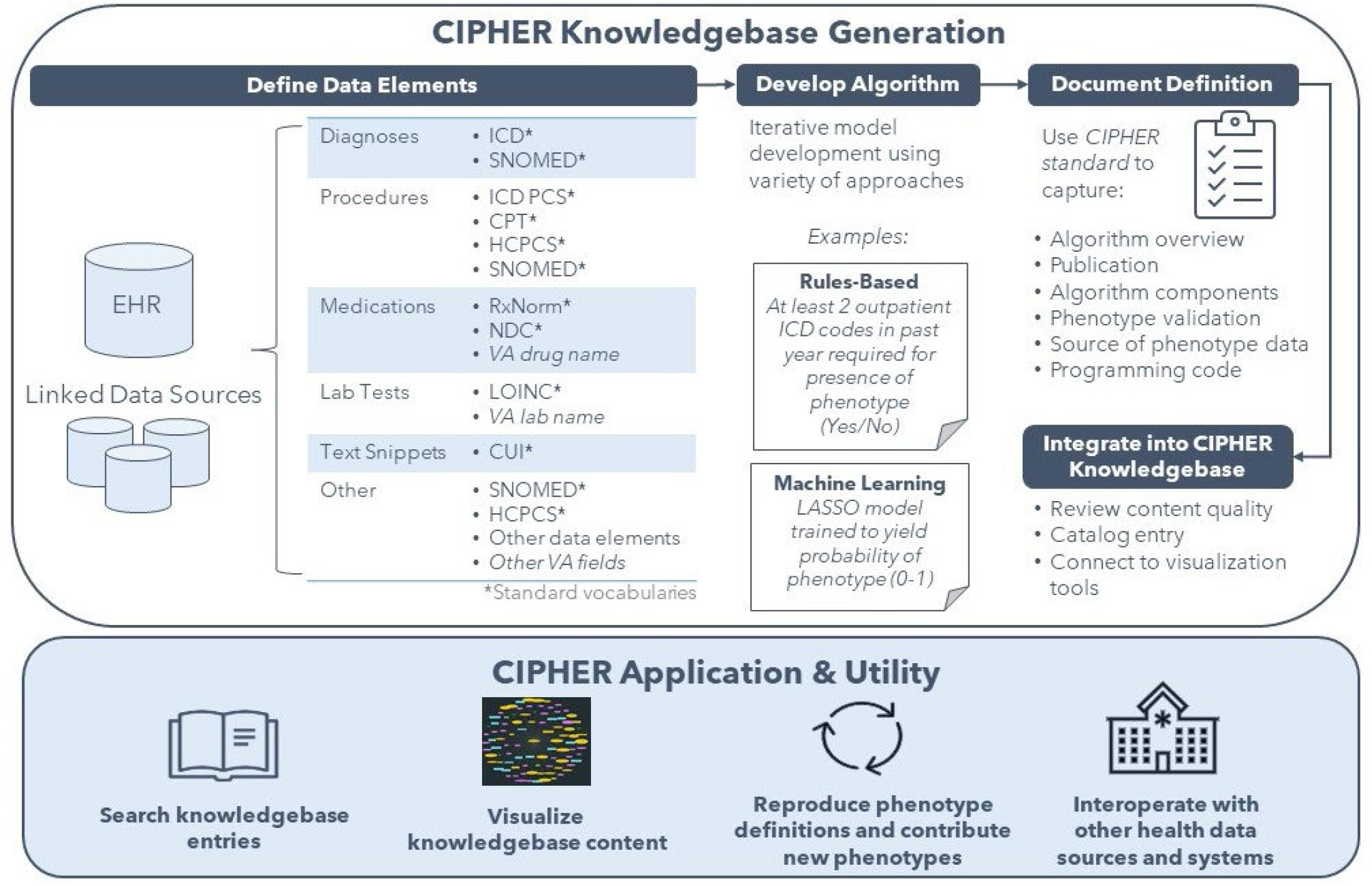
CIPHER Knowledgebase Workflow CPT: Current Procedural Terminology; CUI: Concept Unique Identifier; EHR: Electronic Health Records; HCPCS: Healthcare Common Procedure Coding System; ICD: International Classification of Diseases; LOINC: Logical Observation Identifiers Names and Codes; NDC: National Drug Code. The CIPHER knowledgebase intakes phenotype definitions from contributors using the metadata collection standard and reviews content for quality before publishing to the knowledgebase for community use.

The challenges of phenotyping are not unique to the VA and several phenotype libraries have been created by various research groups as knowledgebases for phenotype definitions. The Phenotype KnowledgeBase (PheKB) hosts phenotype definitions from multiple Electronic MEdical Records and GEnomics (eMERGE) Network sites in the United States.^7^ The HDR UK Phenotype Library is focused on collection of phenotypes from the United Kingdom (UK) EHR.^8^ These libraries demonstrate that phenotyping algorithms can be centralized and cataloged for reuse. However, current metadata collection approaches need a more systematic metadata capture that provides richer context for users and improves transportability across health systems. Through VA’s integrated healthcare system, CIPHER has applied and developed standards for metadata collection which set the foundation for the CIPHER knowledgebase and innovation platform.

The objective of this report is to provide the framework of the CIPHER standard for EHR-based phenotype metadata collection based on our experience through its application in the VA national healthcare system.

## MATERIALS AND METHODS

Existing phenotype libraries, guidelines and desiderata from the literature were evaluated to determine the current landscape of standards and identify remaining gaps.^7-12^ Furthermore insights and experience from investigators, data analysts, and other stakeholders from the VA community have been incorporated to refine cataloging standards used in CIPHER. The development of the CIPHER standard for phenotype metadata collection took an iterative approach which relied on the following four primary principles:

### Know the Audience

The primary audience for reviewing phenotype metadata includes principal investigators, project managers, clinicians, statisticians and data scientists seeking to leverage existing phenotype definitions. The secondary audience includes stakeholders from healthcare administration and operations such as program managers, center directors and scientific officers to query and access available phenotypes developed within the healthcare system to aid in policy decisions and guidelines. Development of these standards catered to the primary audience who are well versed in phenomics science, but also include high-level information for all audiences.

### Provide Context

The scope and purpose of phenotype development must be clearly defined, so that users can determine whether the phenotype algorithm is generalizable for their use case. For example, a definition for diabetes may be created to optimize sensitivity and identify all possible cases of diabetes as an exclusion criterion for a study of new onset diabetes. This is an informative starting point for a case control study seeking high specificity, but further refinement of the definition is advised.

### Facilitate Reproducibility

The standard must provide enough information including data provenance, process, and methods used for a user to replicate the phenotype definition. Reproducibility is key to the utility of collected phenotype metadata. It also enables direct comparison of phenotype prevalence in different settings.

### Enable Adaptability

The standard allows the collection of granular detail to enable reproducibility, but some fields may not be applicable across all phenotypes provided by users. For example, many phenotypes are not formally evaluated for performance, but this is still useful to collect. Additionally, the standard allows the collection of multiple definitions for one phenotype, which capture the nuance between the definitions such as the role of the phenotype in the analysis. Users may then evaluate multiple definitions to determine which one best meets their needs.

## RESULTS

The CIPHER standard for phenotype metadata collection consists of seven domains (Table 1). Each domain consists of standard fields (see *Supplementary Material* for full collection form). Seven of the fields use standard categories used in cataloging.

**Table 1:** CIPHER Phenotype Metadata and Standards for Cataloging

| Metadata Domain | Contents | Standard for phenotype metadata cataloging |  |  |
| --- | --- | --- | --- | --- |
| Phenotype Identification | Unique phenotype name | Free text |  |  |
|  | Abbreviations and keywords | MESH term or free text |  |  |
|  | Phenotype validation status | Working Definition (*in use, but not validated*)<br>Validated Definition (*validated against a standard*) |  |  |
| Algorithm Overview | Data classification | Combat Related | Health Services and Programs | Medications |
|  |  | Demographics | Laboratory Tests | Procedures |
|  |  | Diseases | Lifestyle/Environmental Factors | Vital Signs |
|  |  | Health Access and Metrics |  |  |
|  | Related disease domain | Cardiovascular | Genitourinary | Neurology |
|  |  | Congenital Anomalies | Geriatric | Obstetrics/ Gynecology |
|  |  | Dental | Hematology | Oncology/ Neoplasms |
|  |  | Dermatology | Infectious Disease | Respiratory |
|  |  | Endocrine/Metabolic | Injuries/ Poisonings | Rheumatology |
|  |  | ENT/Ophthalmology | Mental/ Behavioral Health | Symptoms |
|  |  | Gastrointestinal | Musculoskeletal | Women's Health |
|  | Algorithm description (*high level*) | Free text |  |  |
|  | Method used (*phenotyping approach*) | Rules-Based | Machine learning - Unsupervised | Other |
|  |  | Machine learning - Supervised | Machine learning - Other |  |
|  |  | Machine learning - Semi-Supervised |  |  |
|  | Population in which phenotype was developed | Free text |  |  |
|  | Author and contact information | Free text |  |  |
|  | Date created | Date |  |  |
| Acknowledgment and Publication | Publication | Citation and hyperlink |  |  |
|  | Acknowledgment | Free text |  |  |
| Algorithm Components | Algorithm component list and directions for use | ICD-9/10 Codes | Medications |  |
|  |  | ICD-9/10 Procedure Codes | Lab Tests |  |
|  |  | CPT Procedure Codes | Text snippets |  |
|  |  | Clinic Stop Codes | Other |  |
|  | Description of approach and rationale | Free text |  |  |
| Validation | Description of validation | Free text |  |  |
|  | Performance metrics | Sensitivity | PPV |  |
|  |  | Specificity | AUC |  |
|  |  | NPV | Other |  |

| Source of<br>Phenotype Data | Data source | Free text |  |  |
| --- | --- | --- | --- | --- |
|  | Role of phenotype in analysis | Primary Outcome/Exposure<br>Secondary Outcome/ Exposure | Inclusion/Exclusion Requirement<br>Comorbidity/Covariate | Other |
| Additional<br>Information | • Programming code or public code repository link<br>• Tables, figures, slides and associated files<br>• Prevalence data | Free text<br>Images<br>Attachments |  |  |

AUC: Area Under the ROC Curve; CPT: Current Procedural Terminology; ICD: International Classification of Diseases; NPV: Negative Predictive Value; PPV: Positive Predictive Value

The domains of the CIPHER standard are described below:

### Phenotype Identification

Phenotypes are uniquely identified using the convention “*Phenotype Name, subtype (Author)”*. Abbreviations and keywords including Medical Subject Headings terms are collected to facilitate cataloging. Each phenotype receives a status of “Working”, a completed and ready-to-use phenotype, or “Validated”, a phenotype validated via chart review, replication of known associations or another standard.

### Algorithm Overview

This section is intended to provide a snapshot to the reader of the key algorithm information. The “Data Classification” and “Related Disease Domain” fields are used to catalog phenotypes and include categories specific to the VA population such as “Combat Related”. A brief, high-level summary of the algorithm is contained here and is designed to be accessible to any reader. The method used by the algorithm is stated so that the user can quickly determine its complexity. The description of the population used to develop the algorithm informs the user whether the algorithm is generalizable to their population of interest. The date of algorithm creation is included to denote version and the author contact is also provided.

### Acknowledgement and Publication

If the algorithm is affiliated with a published manuscript, the PubMed or preprint journal link is provided. In the absence of a citation, the author will provide an acknowledgment so that work may be attributed to the author by future users.

### Algorithm Components

This domain contains a more descriptive explanation of the data elements used to construct the algorithm and how to use them. The CIPHER standard lists commonly used algorithm components including diagnosis codes, procedure codes, lab tests and medications, but other data elements may be included as well. The phenotype author provides the code list for the definition and describes any inclusion, exclusion, frequency or other requirements for each code set. A description of the entire algorithm is provided which details how each code set is used to create the final phenotype definition. The author also provides rationale for the use of the approach if it is not available in a published manuscript.

### Validation

This section describes the validation of the phenotype, if performed, and performance metrics. Validation practices may range widely from using chart review as a gold standard, comparing against patient-reported data, or replicating a known association [such as replicating expected results from a genome wide association study (GWAS)]. Standard fields used for reporting performance are listed in Table 1.

### Source of Phenotype Data

The source data for phenotype development is listed, which may include VA data and other linked sources. The role of the phenotype in the analysis is also captured using standard responses and the collection method accommodates description of phenotype use for other cases, such as healthcare operations.

### Additional Information

We allow the author to share other resources which may provide context and clarity for the user such as prevalence statistics for applied phenotypes, figures or attached files. This section also contains programming code or a link to a public code repository.

The CIPHER standard has been used to collect phenotype metadata for over 5,000 phenotypes from across the VA user community. (Figure 2)

**Figure 2:**
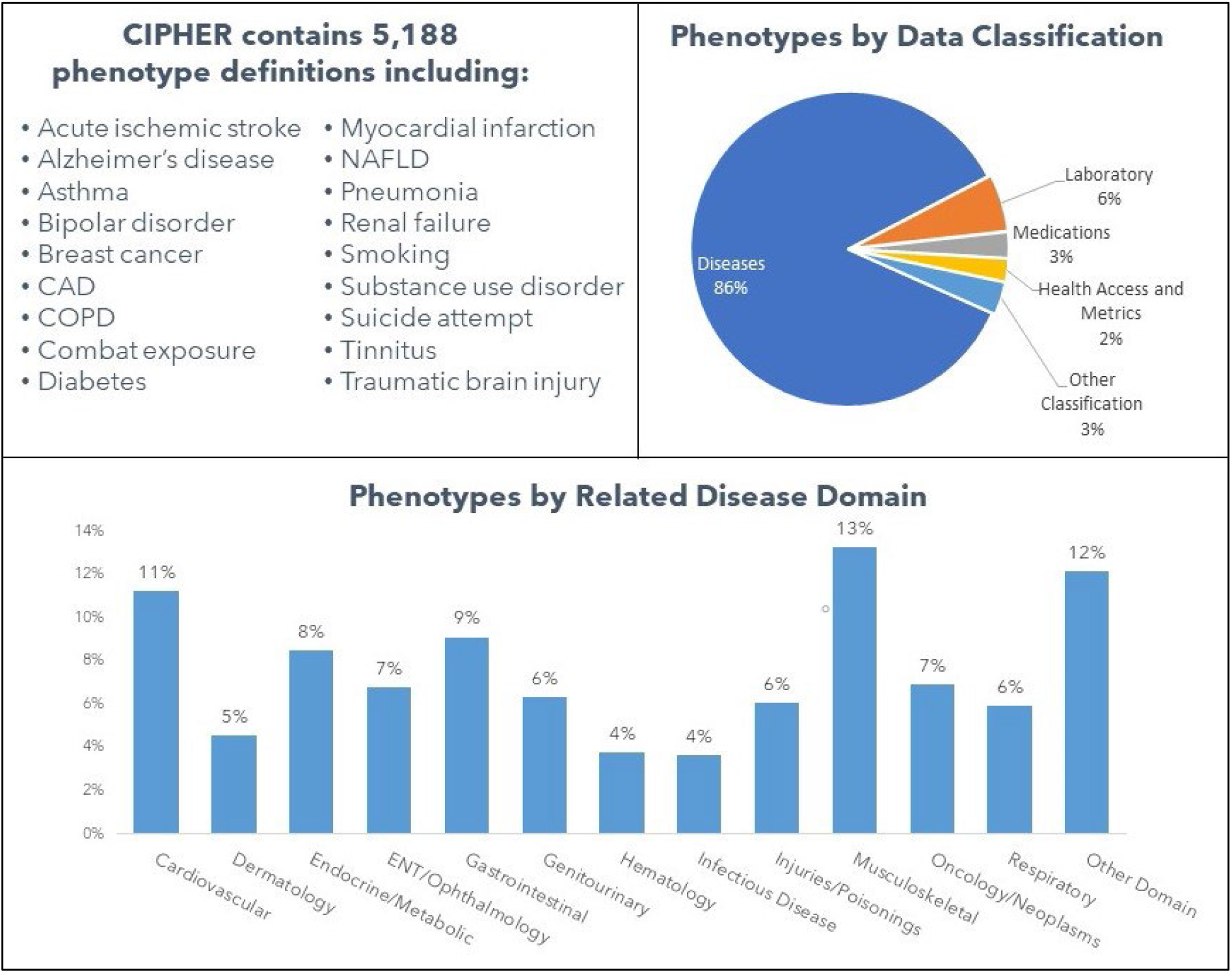
CIPHER Platform Content CAD: Coronary artery disease; COPD: Chronic obstructive pulmonary disease; ENT: Ear, nose and throat; NAFLD: Non-alcoholic fatty liver disease. Other Disease Domains include Congenital Anomalies, Dental, Geriatric, Mental/Behavioral Health, Neurology, Obstetrics/Gynecology, Rheumatology, Symptoms, and Women’s Health. Other Data Classifications include Combat Related, Health Services and Programs, Lifestyle/ Environmental Factors, Procedures and Vitals. Phenotypes may fall into more than one disease domain.

## DISCUSSION

The CIPHER phenotype collection standard is an adaptable metadata collection method that enables reproducibility of EHR-based phenotypes. This standard was iteratively developed with VA community members engaged in phenotype development and provides both detailed information on the phenotype algorithm and a high-level picture of the development and validation process.

Our standard builds on the structure of existing phenotype libraries but requires a more systematic collection of metadata from authors. A comparison against the PheKB and HDR UK phenotype library standards shows the gaps that the CIPHER standard aims to fill. (Table 2) Several fields including overall algorithm description and purpose, phenotyping approach and validation description are irregularly captured in PheKB and not captured in HDR UK. For example, a PheKB page may describe the positive predictive value of a phenotype, but the validation approach is not described. For many data elements a user must read a publication, if available and with sufficient information, to understand the role and method of development for the phenotype. The CIPHER standard aims to showcase this information directly to expedite the user’s understanding of the phenotype and ensure that these fields are systematically captured.

**Table 2:**
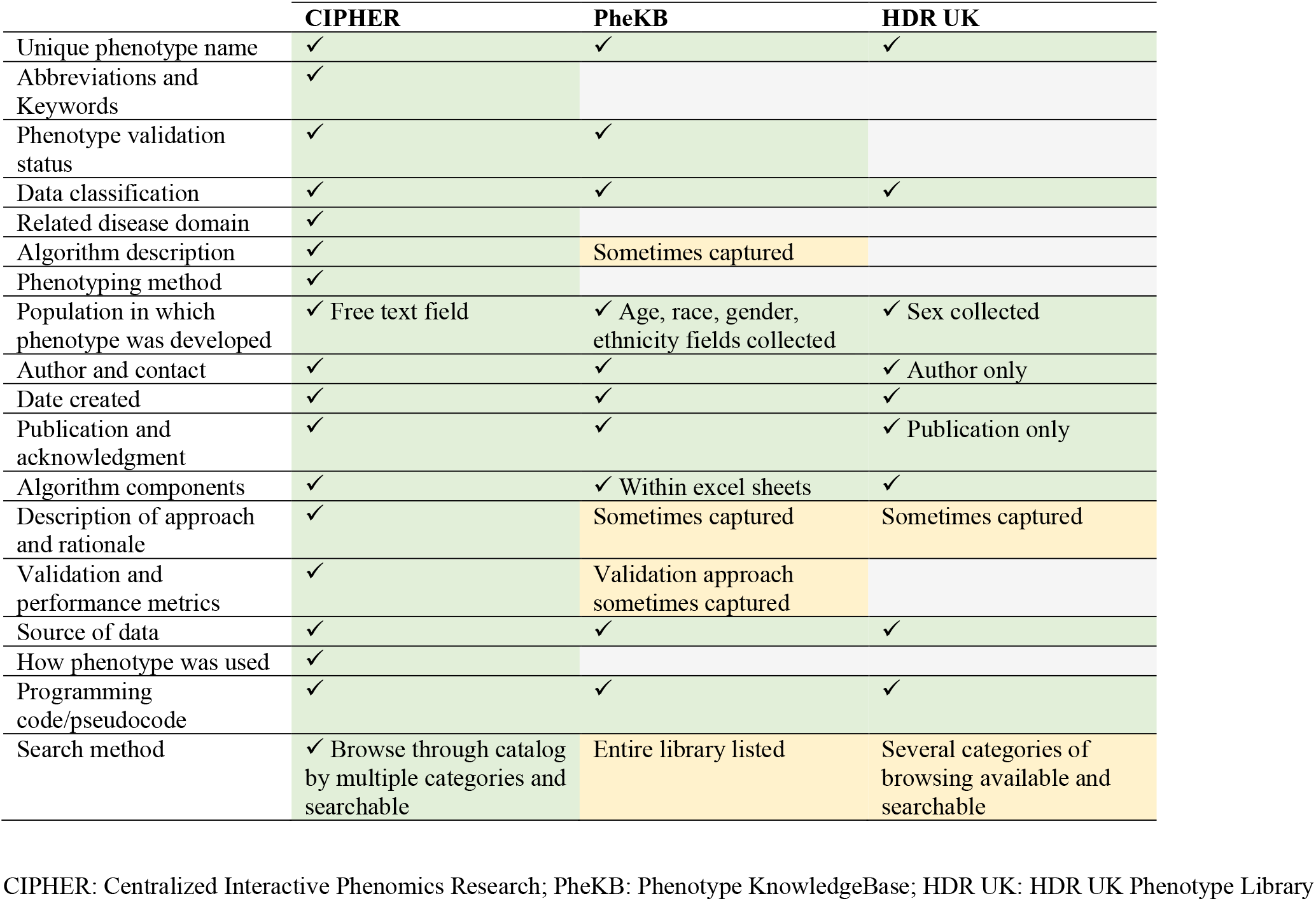
Comparison of Phenotype Libraries

The CIPHER standard enables a centralized capture and dissemination of phenotypes developed by the research and clinical operations communities. (Figure 1) User feedback has been integrated into the development of the CIPHER standard, and ongoing CIPHER quality control and feedback processes improve usability and utility. The earliest version of the CIPHER standard was developed in 2017 to support phenotype collection in the MVP and the MVP scientific community contributed heavily to the iterative development process.^13^ While the CIPHER standard has been implemented in context of the VA EHR, its framework includes standard vocabularies and thus enables the interoperability of the phenotype knowledgebase across various healthcare systems.

The major limitations of the CIPHER standard pertain to its specific use in the VA healthcare system and limited access. While the VA EHR uses standard vocabularies for structured data capture, there are VA-specific data elements that needed to be included in the definition of the standard. The current version of the CIPHER VA phenomics library uses MediaWiki software and is accessible only on the VA internal network. However, CIPHER’s future plans include extending access to the knowledgebase to the wider phenomics community and the public. The CIPHER website will provide a phenotype knowledgebase browsable through smart searching and data visualization tools that display relationships between library concepts, which will provide a significant improvement over currently accessible libraries. The next generation of the library is currently under development and will be accessible at https://phenomics.va.ornl.gov. (Figure 3) CIPHER will continue to highlight phenomics innovation including future content showcasing phenotype metadata used in MVP genetic analyses and resources from the VA Causal Program partnership, among others.

**Figure 3:**
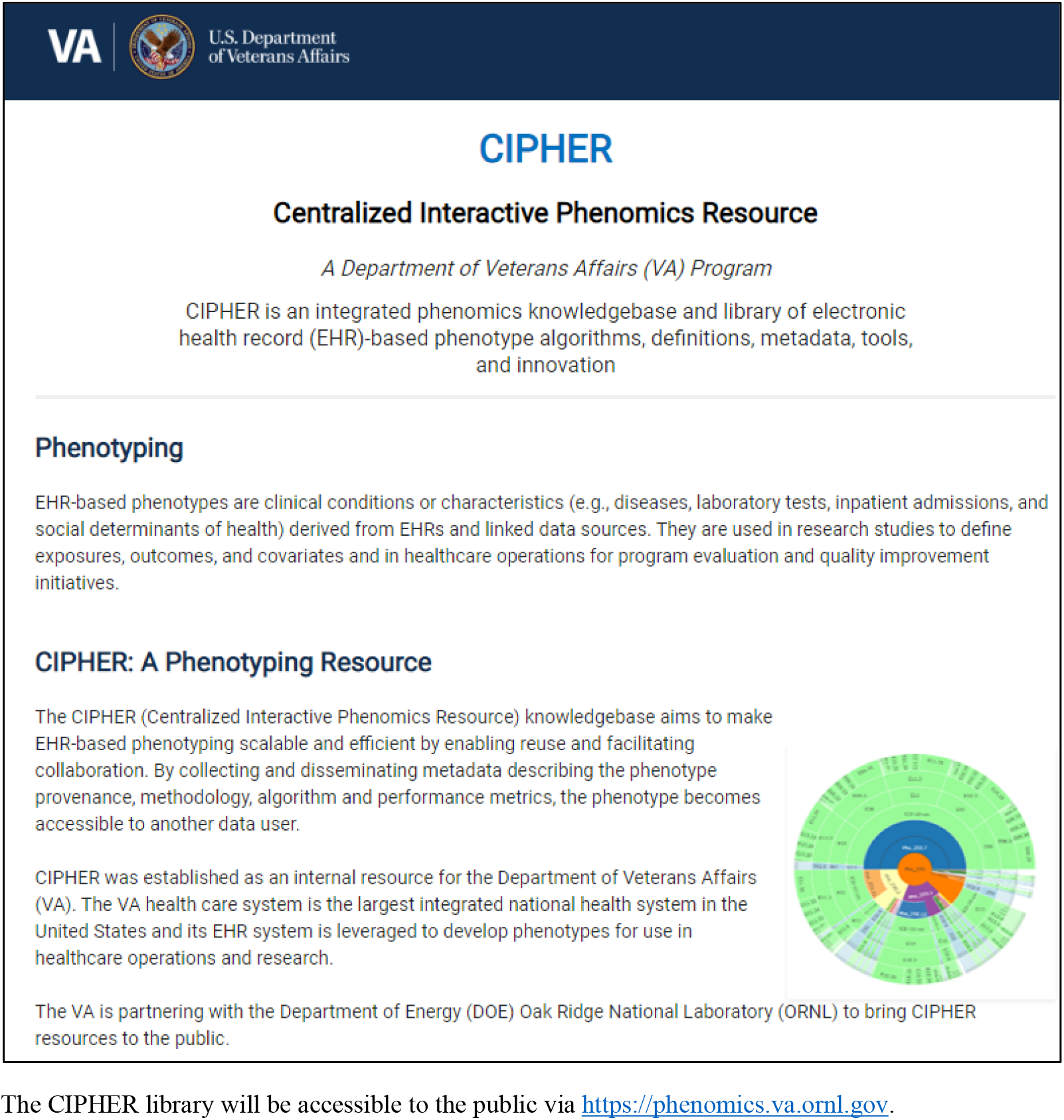
Current CIPHER Website Landing Page The CIPHER library will be accessible to the public via https://phenomics.va.ornl.gov.

## CONCLUSION

The CIPHER standard aims to make EHR-based phenotyping scalable and efficient by enabling reuse and ensuring reproducibility. The standard and its underlying principles for phenotype metadata collection build upon existing phenotype libraries and have been implemented in the VA healthcare system. This standard framework can be applied to other EHR systems and allows interoperability across various systems. CIPHER plans to expand access to its phenotype library beyond the VA and enable all healthcare researchers to utilize this resource.

## Supporting information

CIPHER Phenotype Collection Form

## Data Availability

All data produced in the present work are contained in the manuscript

https://phenomics.va.ornl.gov

