## Supplementary material for "Development of the Centralized Interactive Phenomics Resource (CIPHER) Standard for Electronic Health Data-Based Phenomics Knowledgebase": CIPHER Phenotype Collection Form

### CIPHER - VA Phenomics Library
### Phenotype Entry Form

| Section | Instructions | Phenotype Information |
|---|---|---|
| **Phenotype Name** | Full phenotype name | |
| **Abbreviations and keywords** | Up to 5; this can include MESH terms | |
| **Status (Choose One)** | Choose “Working” if phenotype is completed, but not validated. “Validated” phenotypes are completed and validated via: Gold Standard chart reviews, replication of known associations, etc. | ☐Working<br>☐Validated |
| **Algorithm Overview** | | |
| **Data Classification** | Choose all that apply | ☐Combat Related<br>☐Demographics<br>☐Diseases<br>☐Health Access and Metrics<br>☐Health Services and Programs<br>☐Labs<br>☐Lifestyle/Environmental Factors<br>☐Medications<br>☐Procedures<br>☐Vitals |
| **Related Disease Domain** | Choose all that apply | ☐Cardiovascular<br>☐Congenital Anomalies<br>☐Dental<br>☐Dermatology<br>☐Endocrine/Metabolic<br>☐ENT/ Ophthalmology<br>☐Gastrointestinal<br>☐Genitourinary<br>☐Geriatric<br>☐Hematology<br>☐Infectious Disease<br>☐Injuries/ Poisonings<br>☐Mental/ Behavioral Health<br>☐Musculoskeletal<br>☐Neurology<br>☐Obstetrics/ Gynecology<br>☐Oncology/ Neoplasms<br>☐Respiratory<br>☐Rheumatology<br>☐Symptoms<br>☐Women’s Health |

### CIPHER - VA Phenomics Library Phenotype Entry Form

| | | |
|---|---|---|
| **Algorithm Description** | General summary of algorithm in 1-2 sentences. Include the purpose of the phenotype. | |
| **Population** | Describe population used to develop phenotype or the population this phenotype is applicable to. Include timeframe of data used. Optional: Provide demographics for the cohort used. (Limit: 250 characters) | |
| **Date Algorithm Created** | Date of algorithm release and/or publication | |
| **Author** | Name of project and/or research group/ program that created phenotype | |
| **Contact** | Who is the point person for questions about this phenotype? Add name and email. (Group email is suggested) | |
| **Acknowledgment and Publication** | | |
| **Publication** | Add manuscript citation and/or hyperlink to PubMed or preprint journal, if applicable | |
| **Acknowledgement** | List of authors, grant number and/or acknowledgment language that should be used to cite your work | |
| **Algorithm** | | |
| **Method Used** | Select all that apply | ☐Rules-Based<br>☐ Machine Learning - Supervised<br>☐ Machine Learning - Semi-Supervised<br>☐ Machine Learning - Unsupervised<br>☐ Machine Learning - Other approach<br>☐ Other, *please specify* |

<table>
<tr>
<td>Algorithm Components</td>
<td>Describe the data components used to derive the algorithm and how they were used:<br><br>
<ul>
<li>Description: How the algorithm components are used to develop the phenotype; for example, “at least 2 ICD codes within 6 months”, or “at least 1 ICD code + a positive lab test confirms xx”.</li>
<li>Text snippets refers to components derived from clinical notes which may include structured medical concepts such as CUIs or other extracted data</li>
<li>If natural language processing (NLP) was used to generate the phenotype, please specify the tools, dictionaries and data domains used</li>
<li>For Code List, reference CDW table names where possible to facilitate use of this phenotype.</li>
<li>Note whether there are codes that should be excluded in the description box to the right of the component.</li>
<li>Attach programming codes (SQL, SAS, etc.) used to generate algorithm or link to public repository</li>
<li>Note that CIPHER has established a code repository if you would prefer us to host your code</li>
</ul>
</td>
<td>Description:<br><br>
<table>
<tr><th>Component</th><th>Code List</th><th>Description</th></tr>
<tr><td>ICD-9/10 Codes</td><td></td><td></td></tr>
<tr><td>ICD-9/10 Procedure Codes</td><td></td><td></td></tr>
<tr><td>CPT Procedure Codes</td><td></td><td></td></tr>
<tr><td>Clinic Stop Codes</td><td></td><td></td></tr>
<tr><td>Medications</td><td></td><td></td></tr>
<tr><td>Lab Tests</td><td></td><td></td></tr>
<tr><td>Text snippets</td><td></td><td></td></tr>
<tr><td>Other</td><td></td><td></td></tr>
</table>
</td>
</tr>
<tr>
<td colspan="3">Validation</td>
</tr>
<tr>
<td>Algorithm Validation</td>
<td>Have you performed algorithm validation using chart review or comparison to another standard?</td>
<td></td>
</tr>
</table>

### CIPHER - VA Phenomics Library Phenotype Entry Form

<table>
<tr><td>Description of Validation</td><td>Describe method of validation and process used in 1-2 sentences</td><td></td></tr>
<tr><td>Algorithm Performance Measures</td><td>Choose all that apply. Write the value of the measure to the right.</td><td>☐Sensitivity:<br>☐Specificity:<br>☐NPV (Negative Predictive Value):<br>☐PPV (Positive Predictive Value):<br>☐AUC (Area Under the ROC Curve):<br>☐Other:</td></tr>
<tr><td colspan="3">Source of Phenotype Data</td></tr>
<tr><td>Data Sources Used</td><td>What databases were used to generate the phenotype? Choose all that apply.</td><td>☐CDW (Corporate Data Warehouse)<br>☐CMS (Medicare & Medicaid)<br>☐NDI (National Death Index)<br>☐DoD (Department of Defense)<br>☐MVP (Million Veteran Program)<br>☐COVID SDR (COVID Shared Data Resource)<br>☐OMOP (Observational Medical Outcomes Partnership)<br>☐Other:</td></tr>
<tr><td>Role of Phenotype in Analysis</td><td>Describe how the phenotype was used. Choose all that apply</td><td>☐Primary Outcome/Exposure<br>☐Secondary Outcome/ Exposure<br>☐Inclusion/Exclusion Requirement<br>☐Comorbidity/Covariate<br>☐Other:</td></tr>
<tr><td colspan="3">Additional Information</td></tr>
<tr><td colspan="2">Other information such as phenotype prevalence or useful links, etc.</td><td></td></tr>
<tr><td colspan="3">Attachments</td></tr>
<tr><td colspan="3">Attach any useful documents that you would like to share with library users. If you are including programming code, please ensure that it does not contain any PII/PHI (this includes removal of ANY patient identifier values from your programming code)</td></tr>
</table>
